# Social Disadvantage, Politics, and SARS-CoV-2 Trends: A County-Level Analysis of United States Data

**DOI:** 10.1101/2020.07.11.20151647

**Authors:** Ahmad Mourad, Nicholas A. Turner, Arthur W. Baker, Nwora Lance Okeke, Shanti Narayanasamy, Robert Rolfe, John J. Engemann, Gary M. Cox, Jason E. Stout

**Author notes:** Corresponding author: Jason E. Stout, MD, MHS, Address: Box 102359-DUMC, Durham, NC 27710.

## Abstract

**Background:** Understanding the epidemiology of SARS-CoV-2 is essential for public health control efforts. Social, demographic, and political characteristics at the US county level might be associated with the trajectories of SARS-CoV-2 case incidence.

**Objective:** To understand how underlying social, demographic, and political characteristics at the US county level might be associated with the trajectories of SARS-CoV-2 case incidence.

**Design:** Retrospective analysis of the trajectory of reported SARS-CoV-2 case counts at the US county level during June 1, 2020 – June 30,2020 and social, demographic, and political characteristics of the county.

**Setting:** United States.

**Participants:** Reported SARS-CoV-2 cases.

**Exposures:** Metropolitan designation, Social Deprivation Index (SDI), 2016 Republican Presidential Candidate Victory.

**Main Outcomes and Measures:** SARS-CoV-2 case incidence.

**Results:** 1023/3142 US counties were included in the analysis. 678 (66.3%) had increasing SARS-CoV-2 case counts between June 1 – June 30, 2020. In univariate analysis, counties with increasing case counts had a significantly higher SDI (median 48, IQR 24 – 72) than counties with non-increasing case counts (median 40, IQR 19 – 66; p=0.009). In the multivariable model, metropolitan areas of 250,000 – 1 million population, higher percentage of Black residents and a 10-point or greater Republican victory were independently associated with increasing case counts.

**Limitations:** The data examines county-level voting patterns and does not account for individual voting behavior, subjecting this work to the potential for ecologic fallacy.

**Conclusion:** Increasing case counts of SARS-CoV-2 in the US are likely driven by a combination of social disadvantage, social networks, and behavioral factors. Addressing social disadvantage and differential belief systems that may correspond with political alignment will be essential for pandemic control.

## Background

The SARS-CoV-2 pandemic has been associated with massive societal upheaval worldwide and in the United States (US). In the absence of a vaccine or effective prophylactic medications, the most effective control measures have relied on behavioral change, primarily consisting of social distancing and wearing masks (1–3). However, the ability of individuals in different communities to effectively adhere to behavioral measures relies on an array of social and community factors such as population density, transportation, and poverty. Furthermore, even when these social and community factors are conducive to behavioral change, the willingness of the population in a given community to adhere to behavioral control measures may differ depending on local political ideology. In the early months of the pandemic in the US, social distancing was variable but fairly widespread across the country, despite marked variability in local SARS-CoV-2 epidemiology and government policies (4). Reported increasing “COVID-skepticism” associated with Republican political affiliation may be associated with changes in incident SARS-CoV-2 cases (5). We sought to understand how county sociodemographic and political characteristics might be associated with SARS-CoV-2 incidence.

## Design and Methods

We performed a retrospective analysis of the relationship between the trajectory of reported SARS-CoV-2 case counts at the US county level during the time period 06/01/2020 – 06/30/2020 and social, demographic, and political characteristics. County-level case data were obtained from the Johns Hopkins Coronavirus Dashboard (downloaded 07/02/2020) (6). Counties with >50 cumulative reported SARS-CoV-2 cases as of 05/01/2020 were included in the analysis. We excluded counties with fewer than 50 cumulative cases at that timepoint because sociodemographic factors are unlikely to impact SARS-CoV-2 epidemiology in a meaningful way if there are a very small number of infected persons in a community. The dependent variable was a binary assessment of whether case counts in that community were increasing during the study period (06/01/2020–06/30/2020), derived by fitting a least-squares linear regression line to the case counts during the study period. Counties with a slope >0 were deemed to have increasing case counts, and counties with a slope ≤0 non-increasing case counts.

The independent sociodemographic variables included metropolitan designation, percentage of residents of Black race, percentage of residents of Hispanic ethnicity, and the Social Deprivation Index (SDI) (7). SDI is a composite measure of area level deprivation based on seven demographic characteristics collected in the American Community Survey (2015 update): percent living in poverty, percent of adults ≥25 with <12 years of education, percent single parent household, percent living in a rented housing unit, percent living in an overcrowded housing unit, percent of households without a car, and percent non-employed adults <65 years of age (8, 9). The SDI ranges from 0–100, with higher values indicating more social deprivation. The independent political variable was a binary indicator of whether the Republican presidential candidate won the 2016 election in that county by ≥10 percentage points (10).

Univariate associations were assessed using the Wilcoxon rank-sum test for continuous variables and the chi-squared test for categorical variables. Multivariable associations were assessed using unconditional logistic regression. All independent variables were simultaneously entered into the unconditional regression model. Collinearity was assessed using variance inflation factors, with a cutoff of <3 indicating acceptable collinearity. We examined two-way interactions between SDI and each of the following: the political variable, race/ethnicity, and metropolitan designation, but none of these interactions were statistically significant (p=0.2), and they were removed from the final model. R version 4.0.0 (R Core Team, Vienna, Austria) using the RStudio interface was used for data analysis.

As all data were publicly available and no individual identifiers were used, this study did not require institutional review board review.

## Results

Of 3,142 US counties, 1025 had >50 cumulative reported SARS-CoV-2 cases as of 05/01/2020; 1023 of these had available election results and were included in the analysis. Of these 1023 counties, 678 (66.3%) had increasing daily case counts and 345 (33.7%) had non-increasing case counts between 06/01/2020–06/30/2020. In univariate analysis, counties with increasing case counts had a significantly higher SDI (median 48, IQR 24 – 72) than counties with non-increasing case counts (median 40, IQR 19 – 66; p=0.009). Counties with increasing case counts were more likely to be designated medium-sized metropolitan areas (population ≤1 million) vs. large metropolitan or nonmetropolitan areas (p<0.001), to have a higher percentage of residents of Black race (p=0.013), and to have voted for the Republican presidential candidate in 2016 by a ≥10-point margin (p=0.044) (Table 1).

**Figure 1.**
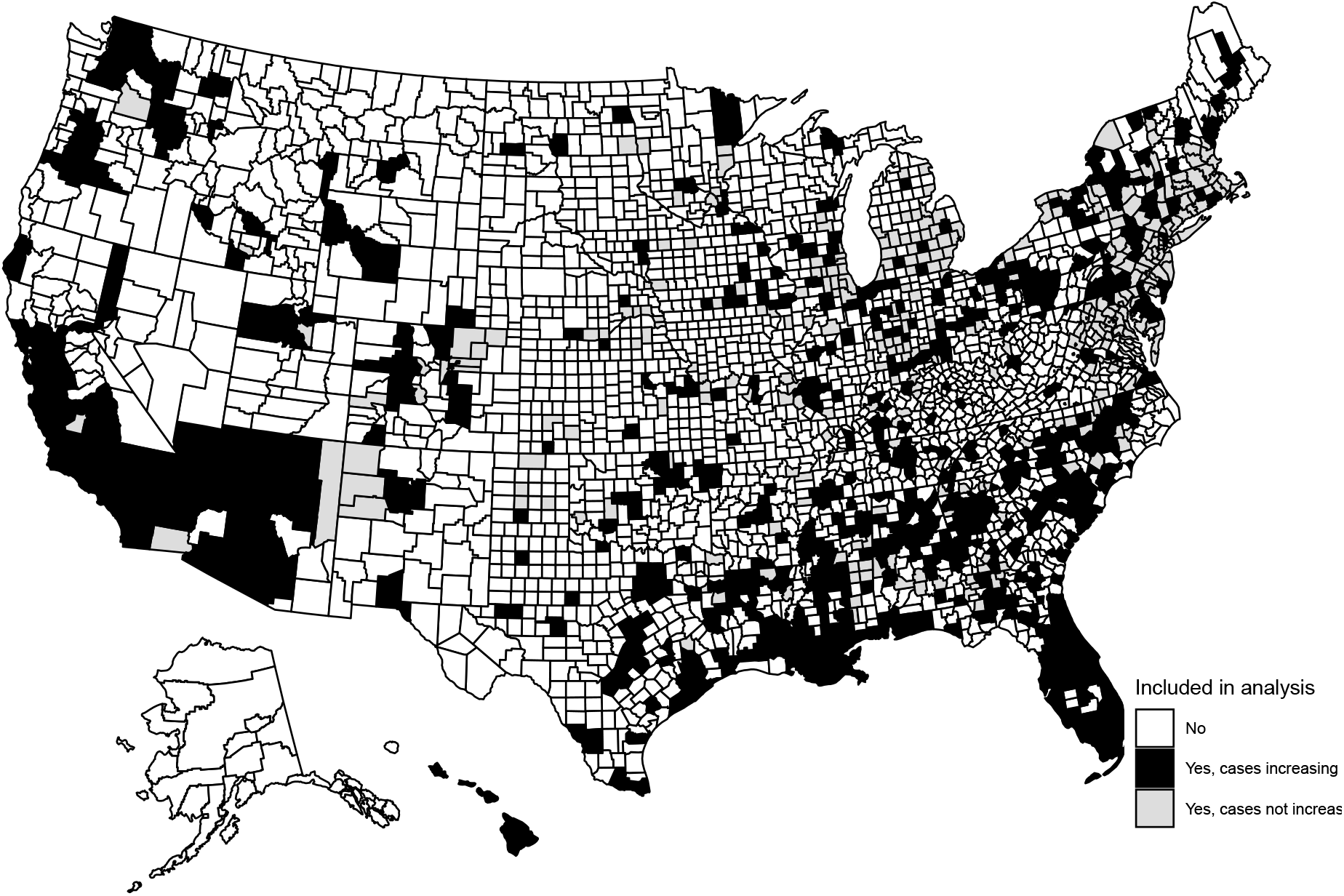
Map of the United States counties included in the analysis.

**Table 1.** Sociodemographic characteristics and SARS-CoV-2 case count trends during the period June 1-June 30,2020 among 1,023 US counties with 50 or more cumulative reported cases as of May 1, 2020.

| Sociodemographic characteristic |  | Increasing case counts (N=678) | Non-increasing case counts (N=345) | p |
| --- | --- | --- | --- | --- |
| Metropolitan designation, n (%) |  | .. | .. | <0.001 |
|  | Metropolitan, 1 million or more population | 193 (28.5) | 118 (34.2) | .. |
|  | Metropolitan, 250,000 – 1 million population | 178 (26.3) | 51 (14.8) | .. |
|  | Metropolitan, < 250,000 population | 130 (19.2) | 53 (15.4) | .. |
|  | Nonmetropolitan | 177 (26.1) | 123 (35.7) | .. |
| % Black race, median (IQR) |  | 9 (3 – 23) | 6 (2 – 17) | 0.013 |
| % Hispanic ethnicity, median (IQR) |  | 6 (3 – 12) | 5 (3 – 12) | 0.856 |
| Social Deprivation Index, median (IQR) |  | 48 (24 – 72) | 40 (19 – 66) | 0.009 |
| Social Deprivation Index by quartile, n (%) |  | .. | .. | .. |
|  | 1 – 17 | 120 (17.7) | 82 (23.8) | .. |
|  | >17 – 39 | 164 (24.2) | 89 (25.8) | .. |
|  | >39 – 60 | 150 (22.1) | 78 (22.6) | .. |
|  | >60 – 100 | 244 (36.0) | 96 (27.8) | .. |
| Republican presidential candidate won by ≥10 percentage points in 2016, n (%) |  | 414 (61.1) | 188 (54.5) | 0.044 |

Table 2 summarizes the univariate and multivariable associations between county characteristics and increasing/non-increasing case counts. In the multivariable model, higher proportion of Black residents, residence in a metropolitan area of 250,000 – 1 million population, and a ≥10-point Republican presidential victory were independently associated with increasing case counts.

**Table 2.** Univariate and multivariable relationships between independent variables and increasing county SARS-CoV-2 case counts June 1-June 30, 2020.

| Sociodemographic characteristic |  | Univariate odds (95% CI) | Multivariable odds (95% CI) | p |
| --- | --- | --- | --- | --- |
| Metropolitan designation, n (%) |  | .. | .. | .. |
|  | Metropolitan, 1 million or more population | Reference | Reference | <0.001 |
|  | Metropolitan, 250,000 – 1 million population | 2.13 (1.45 – 3.14) | 2.04 (1.37 – 3.05) | .. |
|  | Metropolitan, < 250,000 population | 1.50 (1.01 – 2.22) | 1.38 (0.91 – 2.10) | .. |
|  | Nonmetropolitan | 0.88 (0.64 – 1.22) | 0.70 (0.49 – 1.10) | .. |
| % Black race |  | 3.01 (1.29 – 7.01) | 5.73 (1.69 – 19.45) | 0.005 |
| % Hispanic ethnicity |  | 2.16 (0.72 – 6.49) | 3.75 (0.93 – 15.11) | 0.063 |
| Social Deprivation Index by quartile |  | .. | .. | .. |
|  | 1 – 17 | Reference | Reference | .. |
|  | >17 – 39 | 1.26 (0.86 – 1.84) | 1.17 (0.79 – 1.75) | 0.431 |
|  | >39 – 60 | 1.31 (0.89 – 1.94) | 1.02 (0.66 – 1.57) | 0.933 |
|  | >60 – 100 | 1.74 (1.20 – 2.51) | 1.22 (0.73 – 2.04) | 0.456 |
| Republican presidential candidate won by ≥10 percentage points in 2016 |  | 1.31 (1.01 – 1.70) | 1.80 (1.33 – 2.42) | <0.001 |

## Discussion

Our study found that medium-sized metropolitan status, higher proportion of Black residents, and Republican victory in the 2016 presidential elections were independently associated with rising SARS-CoV-2 county case counts during June 2020. This analysis illustrates the complex interplay between demographic and social factors in propagation of the pandemic. In the US, the bulk of cases during the first two months were in urban areas on the East and West coasts, but now the pandemic has taken root in smaller metropolitan areas. The racial disparity in cases persists, perhaps related to a combination of social disadvantage and social networks. However, the finding that voting patterns were independently associated with increased case numbers during June 2020 bears further examination.

One potential explanation involves differential behavior by voting status. During the first few months of the pandemic in the US, voter attitudes toward the pandemic and appropriate behavioral responses were similar across party lines. However, the months of May and June have witnessed increasing partisan divisions in belief and attitudes related to the pandemic and appropriate responses (5, 11). Republican voters are increasingly likely to believe that the severity of the pandemic has been overstated and less likely to believe that public health authorities are providing credible information (11). Furthermore, these beliefs do not seem to be modulated by local case burden (5). Conversely, voting patterns may simply be a surrogate for other public health factors such as relative funding for county health departments or preparedness to perform extensive contact tracing (12).

Our study is limited by the nature of the data as well as the potential for confounding. As our data examines county-level voting patterns, we cannot account for individual voting behavior, subjecting this work to the potential for ecologic fallacy. Our findings may well represent the geographic evolution of the epidemic over time from larger cities to smaller metropolitan areas, with voting patterns merely a surrogate for geographic change. We attempted to adjust for these geographic differences in the analysis, but any such adjustment will incompletely account for temporal trends and other unmeasured confounders.

In conclusion, the evolution of the SARS-CoV-2 pandemic within the U.S. is an illustration of the oft-quoted statement “Social problems, medical consequences.” Increasing cases are driven by both social disadvantage and elevation of personal freedoms over public responsibility. Political polarization is detrimental to effective public health, and as healthcare workers and leaders we must pursue a two-pronged approach of mitigating social disadvantage and appealing to a sense of collective purpose across political boundaries.

## Data Availability

Publically available data

## References

1. Cheng VC, Wong SC, Chuang VW, So SY, Chen JH, Sridhar S, et al. The role of community-wide wearing of face mask for control of coronavirus disease 2019 (COVID-19) epidemic due to SARS-CoV-2. J Infect. 2020;81(1):107–14.

2. Cowling BJ, Ali ST, Ng TWY, Tsang TK, Li JCM, Fong MW, et al. Impact assessment of non-pharmaceutical interventions against coronavirus disease 2019 and influenza in Hong Kong: an observational study. Lancet Public Health. 2020;5(5):e279–e88.

3. Wang Y, Tian H, Zhang L, Zhang M, Guo D, Wu W, et al. Reduction of secondary transmission of SARS-CoV-2 in households by face mask use, disinfection and social distancing: a cohort study in Beijing, China. BMJ Glob Health. 2020;5(5).

4. Badr HS, D. H, Marshall M, Dong E, Squire MM, Gardner LM. Association between mobility patterns and COVID-19 transmission in the USA: a mathematical modelling study. Lancet Infect Dis. 2020.

5. Clinton J, Cohen J, Lapinski J, Trussler M. Partisan Pandemic: How partisanship and public health concerns affect individuals’ social distancing during COVID-19 2020 [updated 6/22/2020. Available from: https://ssrn.com/abstract=3633934.

6. Systems Science and Engineering (CSSE) at Johns Hopkins University (JHU). Coronavirus COVID-19 Global Cases by the Center for Systems Science and Engineering Johns Hopkins University Coronavirus Resource Center website2020 [Available from: https://coronavirus.jhu.edu/map.html.

7. Ingram DD, Franco SJ, National Center for Health Statistics (U.S.). 2013 NCHS urban-rural classification scheme for counties. Hyattsville, Maryland: U.S. Department of Health and Human Services, Centers for Disease Control and Prevention, National Center for Health Statistics; 2014. iv, 73 pages p.

8. Butler DC, Petterson S, Phillips RL, Bazemore AW. Measures of social deprivation that predict health care access and need within a rational area of primary care service delivery. Health Serv Res. 2013;48(2 Pt 1):539–59.

9. Liaw W, Krist AH, Tong ST, Sabo R, Hochheimer C, Rankin J, et al. Living in “Cold Spot” Communities Is Associated with Poor Health and Health Quality. J Am Board Fam Med. 2018;31(3):342–50.

10. McGovern T. US County Level Election Results 08-16 2018 [updated 9/7/2018. Available from: https://github.com/tonmcg/US_County_Level_Election_Results_08-16.

11. Mitchell A, Jurkowitz M, Oliphant J, Shearer E. Three months in, many Americans see exaggeration, conspiracy theories, and partisanship in COVID-19 news: Pew Research Center; 2020 [updated 6/29/2020. Available from: https://www.journalism.org/2020/06/29/three-months-in-many-americans-see-exaggeration-conspiracy-theories-and-partisanship-in-covid-19-news/.

12. Wasfy JH, Stewart C, 3rd, Bhambhani V. County community health associations of net voting shift in the 2016 U.S. presidential election. PLoS One. 2017;12(10):e0185051.

